# Increased Long-Term Mortality in Patients Admitted to the Intensive Care Unit with Health-Care Associated Pneumonia

**DOI:** 10.1101/2021.12.23.21267010

**Authors:** Kristin R. Wise, Jordan A. Kempker, Radu F. Neamu, Ketino Kobaidze

## Abstract

**Background:** Pneumonia is a leading cause of death in the United States. Guidelines for its diagnosis and management are periodically updated, but long-term mortality in critically ill patients has not been well studied.

**Objective:** To determine differences in one-year mortality between patients with community-acquired pneumonia (CAP) and healthcare-associated pneumonia or hospital-acquired pneumonia (HCAP/HAP).

**Methods:** A retrospective multivariate analysis of a prospective cohort study that included all patients admitted to a single-center medical intensive care unit (ICU) from October 1, 2007, through September 30, 2008, with a diagnosis of pneumonia.

**Results:** There were 181 patients admitted to the medical ICU with a diagnosis of pneumonia, 58.0% with HCAP/HAP and 42.0% with CAP. Those with HCAP/HAP had an older age distribution and higher proportions of cardiovascular (79.1% vs. 63.2%, P=0.02) and neurological (36.2% vs. 18.4%, P=0.01) comorbidities. The HCAP/HAP patients demonstrated an increased risk of death within one a year in the unadjusted analysis (HR 1.6, 95% CI 1.1-2.2, P=0.01) that did not remain significant in the multivariate analysis (HR 1.3, 95% CI 0.80-2.10. P=0.29) when adjusting for simplified acute physiology score (SAPS) II score, age category, source of admission and a history of diabetes mellitus, neurological disease or malignancy.

**Conclusion:** Compared to patients admitted to a medical ICU with CAP, those with HCAP/HAP had a higher one-year mortality that is accounted for by the increased co-morbidities associated with a HCAP/HAP diagnosis.

**Summary of Key Points:**

- The American Thoracic Society (ATS) and the Infectious Diseases Society of America (IDSA) developed and periodically update guidelines for the diagnosis and management of community-acquired and nosocomial pneumonia based on the patient-care setting in which pneumonia evolved. ATS/IDSA provides guidelines for empiric antibiotic choices based on the category of pneumonia that is diagnosed.
- Pneumonia is a significant cause of mortality in the United States and when combined with influenza ranks as the eighth leading cause of death nationwide yet little is known about the mortality of critically ill patients with pneumonia that require admission to a medical intensive care unit (ICU).
- Our findings suggest that older age, higher severity of illness at ICU admission, and chronic comorbid illnesses are the main contributors to long-term mortality from pneumonia requiring ICU admission.
- In this cohort, we found an independent association between increased mortality and admission from the general hospital ward rather than directly from the emergency department.
- Our study did not demonstrate that initial guideline-based antibiotic therapy was associated with a reduction in short-term mortality; however, it did demonstrate a high prevalence of resistant pathogens in HCAP/HAP patients, which reflects ATS/IDSA guideline expectations.

## INTRODUCTION

Pneumonia is a significant cause of mortality in the United States (US) and when combined with influenza ranks as the eighth leading cause of death nationwide, causing an estimated 50,000 deaths a year (1). In an effort to confront this public health concern, the American Thoracic Society (ATS) and the Infectious Diseases Society of America (IDSA) developed and periodically update guidelines for the diagnosis and management of community-acquired and nosocomial pneumonias. In 2005, these guidelines outlined a different approach to what was previously known as nosocomial pneumonia, dividing it into three categories based on the patient-care setting in which the pneumonia evolved (2). These three categories are healthcare-associated pneumonia (HCAP), hospital acquired pneumonia (HAP) and ventilator-associated pneumonia. Similarly, specific guidelines were published in 2007 to help clinicians with the management of community acquired pneumonia (CAP) (3).

Since publication, these guidelines have spurred research into the efficacy and safety of their treatment recommendations for each pneumonia class as well as their ability to identify patients at risk for drug-resistant pathogens and higher mortality (4-26). In regards to the latter, six studies have examined mortality differences among the different pneumonia types in hospitalized patients and have shown heterogeneous results (8, 11, 12, 14, 18, 22). Among these studies, there is a paucity of data specifically examining critically ill patients and long-term mortality. To our knowledge, only one recent study has examined this question, demonstrating that pneumonia type for patients admitted to the intensive care unit (ICU) was associated with long term mortality but not short term mortality (12). Given the high incidence of this morbid disease, further information is needed regarding the outcomes of the of the various types of pneumonias presenting to the ICU to better identify the burden of disease they each represent to the community and to help prognosticate for individual patients. In this study, we compared the one-year survival among patients admitted to the medical ICU with CAP, HCAP, and HAP and examined the patient characteristics that are associated with mortality.

## MATERIALS AND METHODS

### Study population

This study is a retrospective analysis of a previously collected prospective cohort of patients admitted to the medical ICU from October 1, 2007, through September 30, 2008, at a single center 511-bed urban, an academic hospital in Atlanta, Georgia. The details regarding the patient population in the above-mentioned database are described elsewhere (27). The study proposal was approved by the Emory University Institutional Review Board.

All adult patients admitted to the ICU within the above period with a diagnosis of pneumonia were included in the study. We excluded patients admitted to the medical ICU with ventilator-associated pneumonia (which generally encompassed patients with tracheostomies from ventilator weaning facilities) and those who did not meet criteria for pneumonia based on documented evidence of a new radiographic infiltrate plus documentation of at least two of the following signs or symptoms: 1) white blood cell count >10,000/ml or <4,000/ml; 2) temperature > 38.3° or < 35° Celsius; 3) purulent secretions from the lower respiratory tract; or 4) new-onset or worsening of dyspnea, oxygen requirements or altered mental status.

### Data Sources and Variables

All data were gathered from the electronic medical record by study investigators and entered into an Excel spreadsheet (Microsoft Corporation). The primary variable of interest was the pneumonia type on admission to the medical ICU. Diagnoses of CAP, HCAP, and HAP were determined by study investigators based on documented evidence of criteria according to the 2005 ATS/IDSA guideline definitions (2). Furthermore, the diagnosis of HAP required the documented absence of an infiltrates on a chest x-ray at the initial hospital admission with the development of a new infiltrate upon transfer to the medical ICU. The primary outcome of interest was one-year all-cause mortality, which was determined by linking patients to their corresponding death records with the US Social Security database.

Patient characteristics collected from the electronic medical record included age, gender, race (self-identified), medical co-morbidities, and source of ICU admission. The severity of illness scores, acute physiology, and chronic health evaluation (APACHE) II and simplified acute physiology score (SAPS) II, were calculated using the standard formula with physiologic variables from the time of the ICU admission. For microbiologic data, we recorded the results of the first blood cultures collected during the period from 6 hours before arrival to the ICU until 3 days after. To capture the etiology for pneumonia, all lower respiratory tract (LRT) cultures obtained during the first 5 days after ICU admission were recorded. An LRT culture was defined as a tracheal aspirate, bronchoalveolar lavage (BAL), bronchial wash, protected brush specimen, or mini-BAL. A multi-drug resistant (MDR) gram-negative pathogen was defined as one with documented decreased susceptibility to at least two of the antibiotic classes recommended for treating pneumonia as delineated by the 2005 ATS/IDSA guidelines (2).

For treatment variables, we recorded all antibiotics administered during the period of 12 hours before arrival in the ICU through ICU discharge. Antibiotic appropriateness was determined by antibiotics given at ICU admission and deemed inappropriate if: 1) they did not include the recommended antibiotic classes for CAP, HCAP, and HAP in the 2005 and 2007 ATS/IDSA guidelines, including double coverage for *Pseudomonas aeruginosa* for HCAP and HAP and atypical coverage for CAP, or 2) they included antibiotics outside of those recommended for respective pneumonia (i.e., using HCAP coverage for CAP). Other treatment variables collected include the use and duration of noninvasive or invasive mechanical ventilation. Outcomes recorded included ICU and hospital length of stay and mortality.

#### Statistical Analysis

All data analyses were performed using SAS 9.3 (SAS Institute Inc. Cary, NC) statistical software. We initially intended to compare CAP, HAP, and HCAP as separate categories, but only 15 (8.0%) subjects had HAP. Therefore, HAP and HCAP were combined for analysis and are referred to as HCAP/HAP in our study. For the bivariate analyses, categorical variables were compared with a chi-square test or Fisher’s exact test. Continuous variables with a normal distribution were compared with a two-tailed t-test. Continuous variables with a non-normal distribution were compared using the Wilcoxon Ranked Sum test. The model for the multivariate analysis was constructed using the covariate selection method described by Hosmer and Lemeshow and is briefly described here (28). Covariates were initially selected for model inclusion if they were associated with both the primary outcome and exposure at a p-value < 0.25 in the bivariate analyses. Then, covariates were retained in the model for a p-value < 0.10 or if upon removal from the model, the pneumonia classification parameter estimate did not change by ≥ 20%. Then each covariate from the entire dataset was entered into this reduced model and retained if they changed the pneumonia classification parameter estimate by ≥ 20%. Final p-values of <0.05 were considered statistically significant.

## RESULTS

### Patient Characteristics

During the study period, 1,357 patients were admitted to the medical ICU of which 181 (13.3%) carried a diagnosis of pneumonia. There were 105 (58.0%) patients with HCAP/HAP and 76 (42.0%) with CAP. Table 1 compares the baseline demographics, preexisting comorbidities, and clinical stay characteristics of CAP and HCAP/HAP patients. When compared to CAP, patients with HCAP/HAP had a significantly older age distribution and higher proportions of cardiovascular (79.1% vs. 63.2%, P=0.02) and neurological (36.2% vs. 18.4%, P=0.01) comorbidities. There were trends towards higher APACHE II and SAPS II scores in HCAP/HAP patients (23.7 vs. 21.8, P=0.07 and 45.4 vs. 41.5, P=0.08, respectively).

**Table 1.** Patient Characteristics by Pneumonia Type in Adults Admitted to a Medical Intensive Care Unit.

| Variable | CAP<br>*n* = 76 | HCAP/HAP<br>*n* = 105 | P value |
| --- | --- | --- | --- |
| **Demographic Characteristics** |  |  |  |
| Age, years, *n* (%) |  |  | 0.01 |
| < 35 | 7 (9.2) | 8 (7.6) |  |
| 35-49 | 21 (27.6) | 11 (10.5) |  |
| 50-64 | 17 (22.4) | 44 (41.9) |  |
| 65-80 | 18 (23.7) | 25 (23.8) |  |
| > 80 | 13 (17.1) | 17 (16.2) |  |
| Gender, female, *n* (%) | 33 (43.4) | 56 (53.3) | 0.19 |
| Race, black, *n* (%) | 54 (71.1) | 79 (75.2) | 0.81 |
| **Preexisting Comorbidities** |  |  |  |
| Past Medical History, *n* (%) |  |  |  |
| Cardiovascular Disease | 48 (63.2) | 83 (79.1) | 0.02 |
| Pulmonary Disease | 24 (31.6) | 45 (42.9) | 0.12 |
| Diabetes Mellitus | 24 (31.6) | 45 (42.9) | 0.12 |
| Chronic Kidney Disease | 17 (22.4) | 32 (30.5) | 0.23 |
| Neurological Disease | 14 (18.4) | 38 (36.2) | 0.01 |
| Malignancy | 12 (15.8) | 18 (17.1) | 0.81 |
| Human Immunodeficiency Virus | 13 (17.1) | 15 (14.3) | 0.60 |
| Gastroenterological Disease | 13 (17.1) | 17 (16.2) | 0.87 |
| Morbid Obesity (BMI ≥40) | 4 (5.3) | 8 (7.6) | 0.53 |
| Immunocompromise | 5 (6.6) | 5 (4.8) | 0.60 |
| **Clinical Stay Characteristics** |  |  |  |
| Location before ICU Transfer, n (%) |  |  | 0.06 |
| Emergency Department | 53 (69.7) | 59 (56.2) |  |
| General Hospital Ward | 23 (30.3) | 46 (43.8) |  |
| APACHE II Score, mean ±SD | 21.8 ± 6.9 | 23.7 ± 7.2 | 0.07 |
| SAPS II Score, mean ±SD | 41.5 ± 15.1 | 45.4 ± 14.3 | 0.08 |
| Appropriate Empiric Antibiotics, n (%) | 29 (38.2) | 18 (17.1) | <0.01 |
| Lower Respiratory Culture Obtained, n (%) | 21 (27.6) | 34 (32.4) | 0.49 |
| MDR Organism Identified, n (%) | 2 (2.6) | 12 (11.4) | 0.05 |
| Endotracheal Intubation, n (%) | 29 (38.1) | 40 (38.1) | 1.0 |
| Noninvasive Ventilation, n (%) | 23 (30.3) | 34 (32.4) | 0.76 |
| ICU Length of Stay, median days (IQR) | 3 (2.0-7.0) | 4 (2.0-8.0) | 0.41 |
| Hospital Length of Stay, median days (IQR) | 9 (5.7 -13.6) | 9.8 (5.3-19.7) | 0.35 |

APACHE, acute physiology and chronic health evaluation; BMI, body mass index; CAP, community-acquired pneumonia; HCAP/HAP, healthcare-associated/hospital-acquired pneumonia; ICU, intensive care unit; IQR, Inter-quartile range; SAPS, simplified acute physiology score; MSSA, Methicillin-sensitive Staphylococcus aureus; MRSA, Methicillin-resistant Staphylococcus aureus; MDR, Multi-drug resistant.

### Main Findings

The one-year survival for critically ill pneumonia patients and their characteristics associated with mortality are shown in Table 2 and their Cox proportional survival curves are depicted in Figure 1. The one-year crude mortality of HCAP/HAP and CAP patients was 62.0% and 34.0%, respectively. HCAP/HAP patients had an increased risk of death within one year in the unadjusted analysis (HR 1.6, 95% CI 1.1-2.2, P=0.01). This finding did not remain statistically significant in the multivariate analysis (HR 1.3, 95% CI 0.8-2.1, P=0.29) when adjusting for SAPS II score, age category, source of ICU admission, and preexisting conditions of diabetes mellitus, neurological disease or malignancy (Table 2). We performed a secondary, exploratory analysis examining mortality at different time points, showing an increase in unadjusted mortality among HCAP/HAP patients at 60 days, but not at 30 or 90 days (Table 3). We ran our multivariable model with the 60-day outcome and again HCAP/HAP did not remain a significant risk factor for mortality.

**Table 2.** Multivariate Cox Proportional Hazards Model for Time from Intensive Care Unit Admission to Death within One Year Among Patients Admitted to a Medical Intensive Care Unit with Pneumonia, N = 181, Likelihood Ratio Test p < 0.0001.

| Variable | Hazard Ratio (95% CI) | P value |
| --- | --- | --- |
| HCAP/HAP vs. CAP | 1.30 (0.80-2.10) | 0.29 |
| SAPS II Score | 1.02 (1.01-1.04) | <0.01 |
| Admitted From General Hospital Ward vs. ED | 1.66 (1.05-2.49) | 0.03 |
| Age Category |  |  |
| < 35 | Referent |  |
| 35-49 | 0.81 (0.28-2.40) | 0.70 |
| 50-64 | 1.04 (0.39-2.80) | 0.94 |
| 65-80 | 1.54 (0.57-4.17) | 0.39 |
| > 80 | 1.92 (0.69-5.39) | 0.21 |
| Preexisting Comorbidities |  |  |
| Neurological Disease | 0.98 (0.59-1.62) | 0.94 |
| Diabetes Mellitus | 0.72 (0.40-1.12) | 0.14 |
| Malignancy | 2.70 (1.61-4.51) | <0.01 |

CAP, community-acquired pneumonia; ED, emergency department; HCAP/HAP, healthcare-associated/hospital-acquired pneumonia; SAPS, simplified acute physiology score.

**Figure 1.**
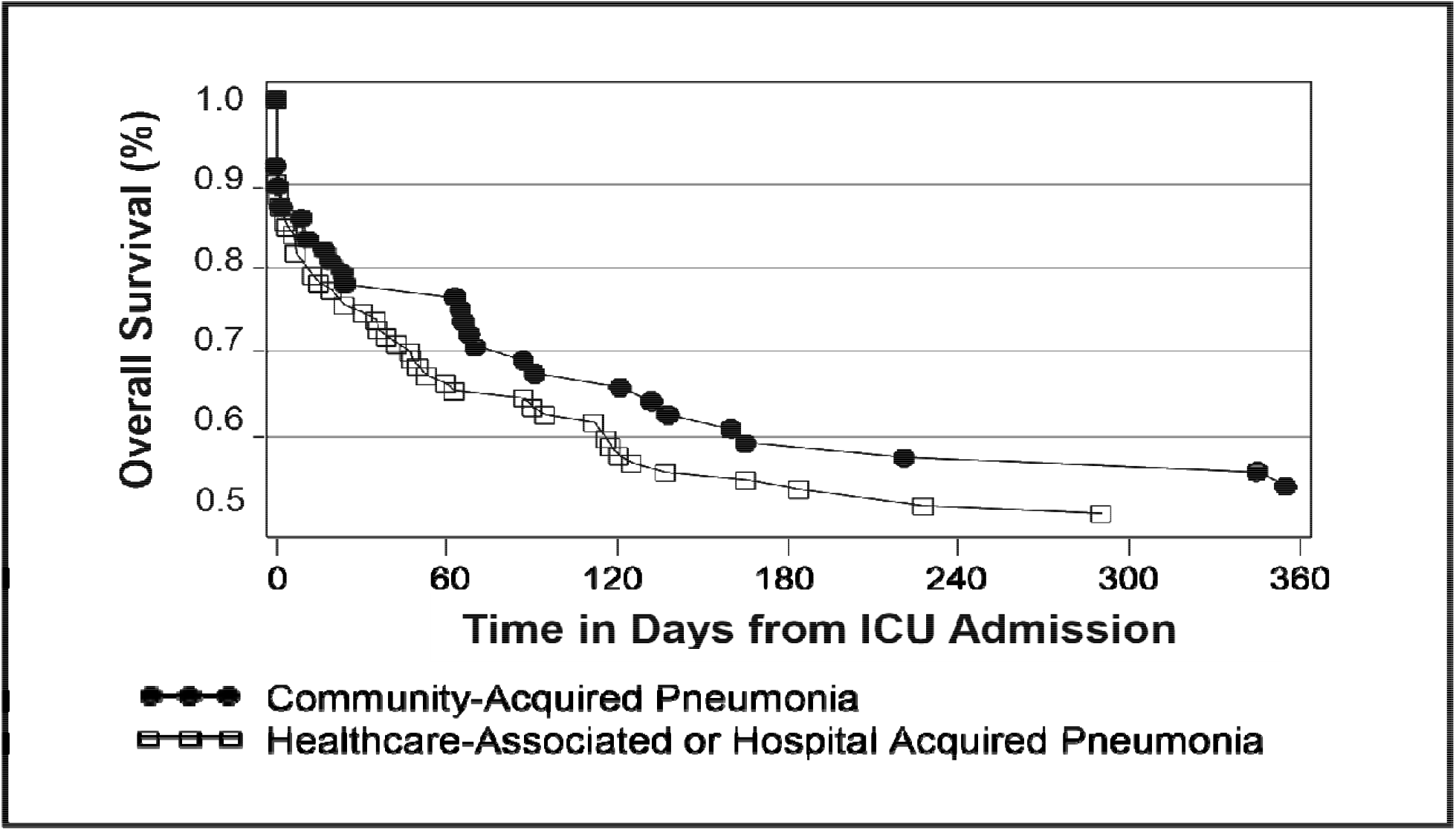
Cox Proportional Survival Curves for Mortality From Intensive Care Unit (ICU) Admission to One Year Among Patients with Pneumonia Admitted to a Medical ICU. Stratified by Pneumonia Type and Adjusted for Simplified Acute Physiology Score (SAPS) II, Location Before Admission, Age Category and Preexisting of Neurological Disease, Diabetes Mellitus and Malignancy. N = 181.

**Table 3.** Exploratory Analysis of Different Mortality Endpoints in Patients with Pneumonia Admitted to the Intensive Care Unit by Pneumonia Type.

| **Mortality Endpoint** | **CAP Mortality**<br>**n (%)** | **HCAP/HAP Mortality**<br>**n (%)** | **Unadjusted RR (95%CI)**<br>**HCAP/HAP v. CAP** | **P value** |
| --- | --- | --- | --- | --- |
| ICU | 7 (9.2) | 14 (13.3) | 1.3 (0.7-2.4) | 0.39 |
| 30-Day | 17 (22.4) | 33 (31.4) | 1.3 (0.9-2.1) | 0.18 |
| 60-Day | 17 (22.4) | 43 (41.0) | 1.7 (1.1-2.7) | 0.09 |
| 90-Day | 23 (30.3) | 46 (43.8) | 1.4 (1.0-2.1) | 0.06 |
| One Year | 32 (42.1) | 64 (61.0) | 1.6 (1.1-2.2) | 0.01 |

CAP, community-acquired pneumonia; HCAP/HAP, healthcare-associated/hospital-acquired pneumonia; ICU, intensive care unit.

### Microbiology of Pneumonias and Antibiotic Treatment

Cultures were not performed in 5.7% and 5.3% of HCAP/HAP and CAP patients, respectively. The first culture obtained was a blood culture in the majority of both HCAP/HAP (84.8%) and CAP (81.6%) patients. Lower respiratory cultures were performed in 32.4% of HCAP/HAP and 27.6% of CAP patients. Figure 2 summarizes the microbiology of the pneumonias in the study group. Blood and respiratory tract cultures remained negative in 52.6% of CAP patients and 43.8% of HCAP/HAP patients. Methicillin-resistant Staphylococcus aureus (MRSA) and MDR gram-negative bacilli were not cultured among the CAP patients while these organisms comprised the infectious agent in 6.7% and 3.8% of the HCAP/HAP patients, respectively.

**Figure 2.**
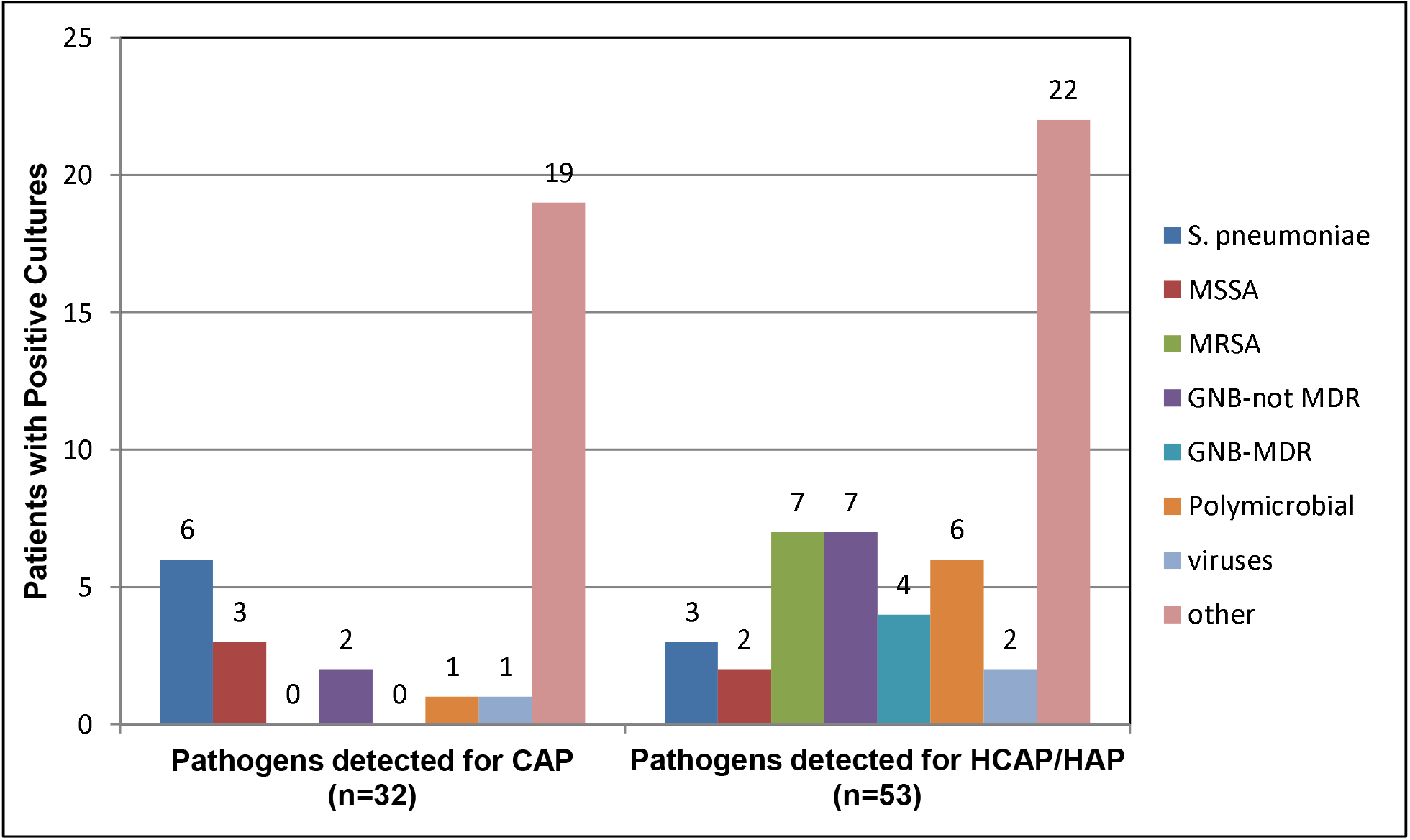
Pathogens Detected in Respiratory and Blood Cultures According to Pneumonia Type. The other category included yeast, nontuberculous mycobacterial species, unidentified gram positive cocci and pneumocystis jirovecii in decreasing order of frequency. CAP, community-acquired pneumonia; HCAP/HAP, healthcare-associated/hospital-acquired pneumonia; MSSA, Methicillin-sensitive Staphylococcus aureus; MRSA, Methicillin-resistant Staphylococcus aureus; MDR, Multi-drug resistant.

When compared to HCAP/HAP patients, those with CAP were more likely to have the appropriate initial antibiotic therapy at admission to the ICU (38.2% vs. 17.1%, P<0.01) as shown in Table 1. However, appropriate antibiotics did not adid not affect one-year mortality, with 27.1% of the survivors and 25.0% of non-survivors receiving appropriate antibiotics (P=0.75).

### Other Outcomes

Invasive mechanical ventilation was required in 38.1% of the CAP patients and 38.1% of the HCAP/HAP patients (Table 1). There were no significant differences in the use of non-invasive positive pressure ventilation or ICU or hospital length of stay between the two pneumonia groups.

## DISCUSSION

This study compares the characteristics and outcomes between patients admitted to a single-center medical ICU with a diagnosis of CAP or HCAP/HAP over one year. Patients with HCAP/HAP were older with more medical comorbidities and more resistant pathogens. While our study identified that the HCAP/HAP patients had an increased, unadjusted one-year mortality when compared to CAP, this finding did not persist when adjusted for SAPS II score, age, source of ICU admission and preexisting diabetes mellitus, neurological disease, or malignancy. These findings suggest that older age, higher severity of illness at ICU admission, and chronic comorbid illnesses are the main contributors to long-term mortality from pneumonias requiring ICU admission.

We compared our mortality findings with the existing literature, which focuses mostly on short-term mortality. To date, we know of only one study that has examined long-term mortality by pneumonia type in patients admitted to ICU. This study demonstrated no difference in adjusted short-term mortality but showed a higher unadjusted one-year mortality among patients with HAP (12). This may be similar to our unadjusted findings of an increased risk of death within one year for HCAP/HAP patients that did not remain statistically significant in our multivariate analysis. While five studies have found that HCAP carries higher unadjusted mortality, this association remained in only one of the three studies that adjusted for confounders. Not surprisingly, these data have also shown that patients with HCAP are older with more severe chronic comorbidities and often present with more severe illness (8, 11, 14, 18, 22). In regards to the effects of comorbid illnesses, one study examining the long-term mortality of patients hospitalized with CAP also found that older age, cerebrovascular disease, and cardiovascular disease were associated with increased mortality (29). Overall, the sum of these data, combined with our study results, suggest that the current pneumonia classification is not an independent risk factor for mortality but rather a marker of more chronic and severe illness.

In addition to our main results, our study identified several other interesting findings. In this cohort, we found an independent association between increased mortality and admission from the general hospital ward rather than directly from the emergency department. A similar association was found in one study that examined the source of ICU admission for patients with septic shock (30). In that study, in comparison to patients admitted from the general ward, those from the emergency department had lower in-hospital mortality, less mechanical ventilation utilization, and shorter times to achieving ScVO_2_ goals (30). As our study was not designed to compare the differences between patients admitted to the medical ICU from these two sources, we can only speculate as to whether treatment in the emergency department was protective or whether admission from the hospital ward was confounded by co-morbid and concurrent illness.

Finally, our study did not demonstrate that initial guideline-based antibiotic therapy was associated with a reduction in short-term mortality; however, it did demonstrate a high prevalence of resistant pathogens in HCAP/HAP patients, which reflects ATS/IDSA guideline expectations. There were no cultures positive for MDR gram-negative bacilli or MRSA among ICU patients with CAP while these organisms were present in 10.5% of HCAP/HAP patients. The existing published literature demonstrates varying conclusions with only a portion of the studies showing a high prevalence of *Pseudomonas aueroginosa* and MRSA in both HCAP and CAP patients (4, 19, 25, 26, 31). Such discrepancies may explain the inconsistent reported results on the association between guideline-based antibiotic therapy and mortality (7, 10, 13, 15-17, 23, 24). Other investigators have sought alternate means to identify those at higher risk of resistant pulmonary pathogens although significant research is still needed in this area (6, 9, 20, 21).

There are limitations to the present study that must be noted. This is a single-center study and as the microbiology of pneumonia may vary according to the setting, our results may not be generalizable. Although the initial cohort was captured prospectively, study investigators determined the diagnoses of pneumonia on retrospective chart review. While the use of specific criteria to define the various pneumonia types increases internal validity, there may be confounding factors in original medical documentation that led to misclassification errors in the study. Furthermore, the retrospective design may have limited an aggressive approach to the microbiologic diagnosis of pneumonia, resulting in a lower proportion of patients with a LRT culture. Despite these limitations, this study continues research in an important area and is able to provide additional data regarding the clinical characteristics, microbiologic findings and mortality outcomes of patients admitted to ICU with a diagnosis of pneumonia obtained from a realistic practice setting.

## CONCLUSION

This study demonstrates higher one-year mortality among patients admitted to a medical ICU with HCAP/HAP compared to those admitted with CAP that is likely accounted for by the increased co-morbidities associated with a HCAP/HAP diagnosis. Further evidence is needed to distinguish the true, and likely evolving, microbiologic relevance of the distinction between HCAP/HAP and CAP to guide future treatment recommendations.

## Data Availability

All data produced in the present study are available upon reasonable request to the authors

## Acknowledgments

The authors have neither financial interests nor conflicts of interest to disclose.

